# Comprehensive genetic screening of the South African Parkinson’s disease study collection using the NeuroBooster array

**DOI:** 10.64898/2025.12.17.25341881

**Authors:** Kathryn Step, Lusanda Madula, Nicole Kuznetsov, Elizna Maasdorp, Jonathan Carr, Riaan van Coller, Ignacio F. Mata, Soraya Bardien, Global Parkinson’s Genetics Program (GP2)

**Author notes:** **Correspondence to:** Soraya Bardien, PhD Division of Molecular Biology and Human Genetics, Faculty of Medicine and Health Sciences, Stellenbosch University, Cape Town, South Africa. These authors share first authorship (Kathryn Step, Lusanda Madula).

## Abstract

Parkinson’s disease (PD) is a complex neurodegenerative disorder with a substantial genetic influence. To better characterize the genetic landscape of PD in South Africa, we conducted the largest genetic screening to date for pathogenic single nucleotide and copy number variations (CNVs) using genotyping array data from 689 PD probands. We identified 16 unique missense variants, confirming 15 with Sanger sequencing, in 47 individuals across seven well-established PD genes, with *GBA1* and *PRKN* being most frequent. Also in known PD genes, 18 variants of unknown significance were found. Additionally, CNV analysis using CNV-Finder revealed seven novel CNVs, five in *PRKN* and two in *SNCA*, of which, six were validated with Multiplex Ligation-dependent Probe Amplification. The findings highlight the contribution of both rare variants and structural rearrangements to PD in this underrepresented population. This study underscores the importance of expanding genetic research in African cohorts to improve global understanding of PD etiology.

## Introduction

Parkinson’s disease (PD) is a common neurodegenerative disorder with a disease etiology linked to both environmental and genetic factors.^1^ Understanding the genetic etiology of PD can clarify the biological pathways involved in disease development, inform risk prediction, and improve strategies for disease prevention, diagnosis, and management.^2^

Approximately 5-10% of individuals with PD carry a pathogenic disease variant in a single gene, representing monogenic forms of the disease.^3,4^ In many cases, monogenic forms of the disease are attributed to highly penetrant and rare genetic variants.^2^ Several genes have been linked to PD, particularly monogenic forms, showing either an autosomal dominant or autosomal recessive mode of inheritance. Notably, of these, six genes (*LRRK2*, *PRKN*, *SNCA*, *PINK1*, *PARK7/DJ-1*, and *VPS35*) have been unequivocally linked to monogenic PD.^4^ Variants in *GBA1*, although not monogenic, remain one of the most significant risk factors for PD.^5^ All seven genes are commonly included in PD diagnostic panels^3,6^.

In addition to missense variants and small indels (short insertions or deletions of DNA), the disease etiology can be attributed to genetic structural variants, resulting in a different number of DNA segment copies than normal.^7^ These are known as copy number variations (CNVs) and include deletions, duplications, insertions, and genomic rearrangements. Although CNVs have been shown to play a role in PD pathogenicity,^8^ they are generally rare and observed in a small proportion of affected individuals at any given gene locus.^9^ Notable CNVs in the context of PD commonly include exonic deletions and duplications in *PRKN* and *PINK1* as well as whole gene duplications and triplications of *SNCA*.^10–12^

Although significant progress has been made in understanding the genetic basis of PD, most studies have focused on European populations, leaving the genetic etiology of PD in African populations, including the South African population, largely uncharacterised. Previous PD research in South Africa employed wet-lab screening techniques, including high-resolution melt analysis, Sanger sequencing, and multiplex ligation-dependent probe amplification (MLPA), to detect pathogenic missense variants, indels, and CNVs commonly observed in populations of predominantly European ancestry. However, screening was limited to a small subset of the study collection and revealed that known PD genetic causes are relatively rare in this cohort.^8,13–15^

To address this gap, we performed a comprehensive genetic screening using genotyping array data for the South African PD Study Collection to identify pathogenic variants including CNVs. We further sought to validate these findings using independent techniques.

## Methods

An overview of the workflow is provided in **Figure 1**.

**Figure 1:**
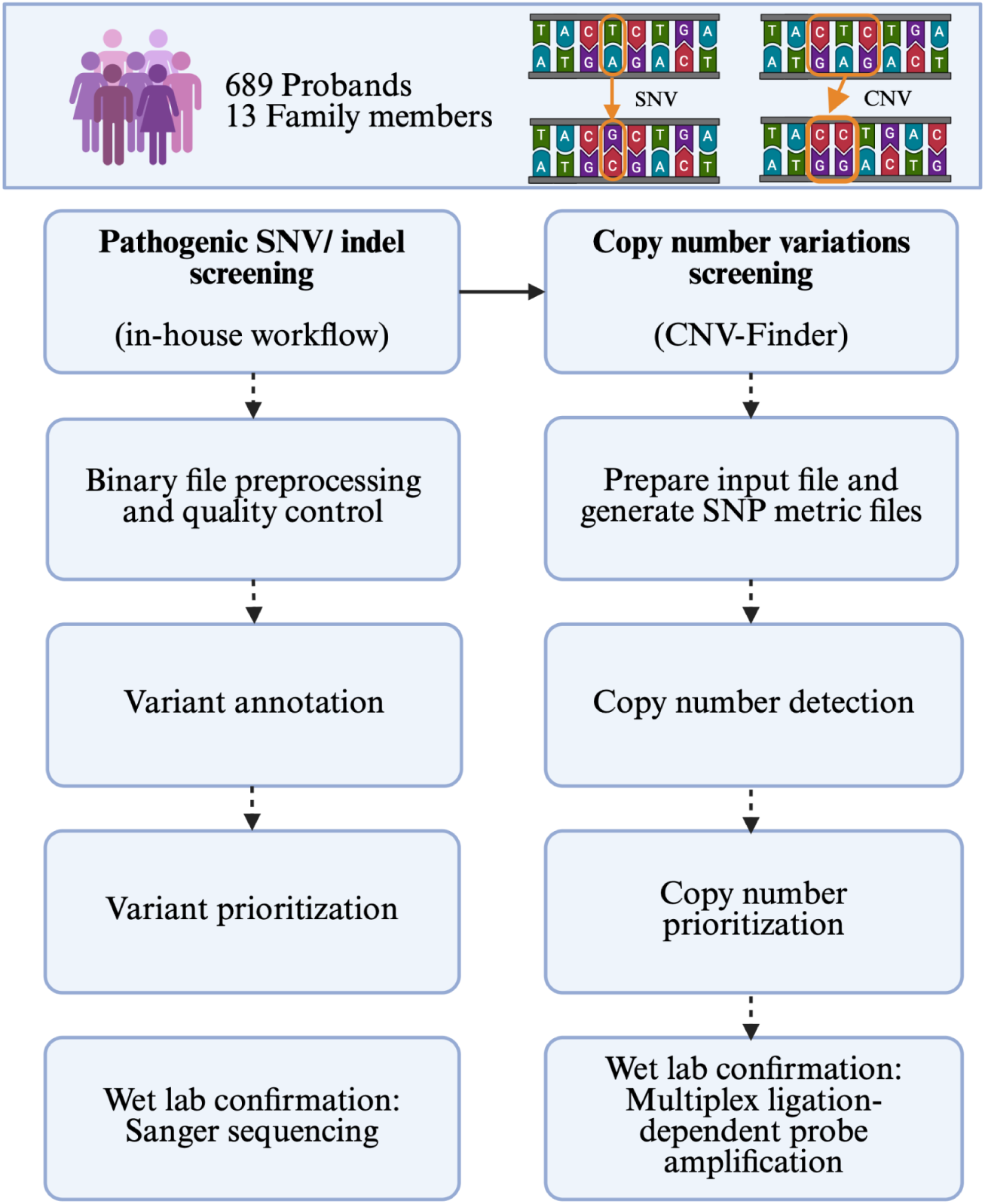
Overview of the study workflow. CNV, copy number variation; SNV, single nucleotide variant.

### Study participants and genotyping

Study participants were recruited over an 18 year period, from 2002 until June 2020,^16^ as part of the South African PD study collection. Individuals living with PD (n=689 probands; **Supplementary Table 1**) were diagnosed using the Queen’s Square Brain Bank Criteria.^17^ Ethical clearance was obtained from the Health Research Ethics Committee at Stellenbosch University, South Africa (2002C/059). The study collection was genotyped using the NeuroBooster array (v1.0, Illumina, San Diego, CA) through a collaboration with the Global Parkinson’s Genetics Program (GP2).^18,19^ Previous research has shown the ancestral makeup of the South African PD study collection is 56% European, 18.8% African, 13% indigenous Nama, 6.9% South Asian, and 5.2% Malay.^20^

### Quality control, file preparation, and processing for single nucleotide variant screening

The present study made use of genotyping data. For the pathogenic single nucleotide variant screening, quality control (QC) was performed using PLINK v1.9.^21^ Data missingness was filtered at 10% in individuals and 5% in variants, individuals with sex discrepancies were removed (female F: 0.5; male F: 0.8), variants that deviated from Hardy-Weinberg equilibrium (controls: 1×10^-6^; cases: 1×10^-10^), and those that were more than 3 standard deviations from the mean heterozygosity rate, were removed.

An in-house computational workflow was developed and used to identify and prioritize pathogenic missense variants and indels. Briefly, this consisted of file preparation and conversion from PLINK binary files to variant call format files. Variants were annotated using ANNOVAR v2020-06-08 with all essential databases in Genome Reference Consortium Human Build 38,^22^ including the cytogenetic bands, genome aggregation, nonsynonymous functional prediction, and clinical variant databases. The analysis was performed in two stages. First, the computational workflow was optimized using a subset of probands (n=13) with five pathogenic missense variants previously identified through wet-lab screening conducted by the South African PD research group (**Supplementary Table 2**). This ensured that the computational workflow could reliably detect known variants. In the second stage, the analysis was restricted to individuals with array genotyping data but without a previously established genetic cause of disease (n=645), allowing for the identification of additional pathogenic missense variants. The ANNOVAR output was aligned to the original files to confirm which individual carried which variant. This was followed by restricting the annotated phenotype to “*Parkinson*” or “*Parkinsonian*”. The files were examined using the following *in-silico* pathogenicity prediction tools and criteria, with variants required to meet at least one of these thresholds to be included: SIFT score < 0.05,^23^ FATHMM score < 0,^24^ PolyPhen-2 score > 0.5,^25^ CADD score >= 20,^26^ MetaLR > 0.8,^27^ and MetaSVM > 0.8.^27^ Finally, variants were filtered using ClinVar (July 2025),^28^ retaining only those classified as “pathogenic”, “likely pathogenic”, or “conflicting pathogenicity” due to their reported associations with the disease. Where DNA from family members was available, co-segregation of the identified pathogenic variants was assessed using Sanger sequencing, as described below. In addition to pathogenic variants, variants of unknown significance (VUS) were also screened using the same workflow. The only modification was adjusting the clinical significance filtering to include variants classified as “uncertain significance” instead of “pathogenic” or “likely pathogenic” according to ClinVar (accessed July 2025).^28^ The prioritized VUS were not confirmed using Sanger sequencing. Additionally, all identified variants in *GBA1* were analysed with GBA1-PD Browser v1.0.^5^

### Validation of single nucleotide variants

The identified putative pathogenic missense variants were confirmed using Sanger sequencing. This included primer design, polymerase chain reaction (PCR) optimization, and Sanger sequencing. The oligonucleotide primers were designed using the Integrated DNA Technologies PrimerQuest™ tool and *in-silico* PCR with the University of California Santa Cruz *in-Silico* PCR (https://genome.ucsc.edu/cgi-bin/hgPcr). Detailed information regarding the primers is included in **Supplementary Table 3**. A gradient PCR was run using the Eppendorf Mastercycler® X50 to optimize the annealing temperatures for the reactions. Following optimization, the PCR reactions were run using the Applied Biosystems GeneAMP® 2700 (**Supplementary Table 4 and 5**). The PCR products were viewed using 2% agarose gel electrophoresis. Due to the presence of the highly homologous *GBA1LP* pseudogene, the aforementioned protocol was modified when validating the variants in *GBA1*. Here, genomic DNA was amplified using long-range PCR targeting a 7kb fragment, which was viewed using a 0.5% agarose gel and exon-specific primers (nested primers), at a higher-than-normal concentration, were used for Sanger sequencing. All Sanger sequencing was done at the Central Analytical Facilities DNA Sequencing Unit, Stellenbosch University (**Supplementary Methods**). The DNA chromatograms were analyzed and interpreted using Molecular Evolutionary Genetics Analysis v12.^29^

### File preparation and implementation of CNV-Finder for copy number variations detection

CNV-Finder was applied to the array data using the same workflow as previously described.^30^ Through prior CNV screening, the South African PD Research Group had identified CNVs in 14 probands and six family members (**Supplementary Table 6**). These previously validated findings were used as reference points to confirm that CNV-Finder was accurately detecting true CNVs in the current dataset. The focus of the screening for the present study was on the probands without a previously identified PD pathogenic variant, including an investigation of potential compound heterozygous CNVs when combined with the single nucleotide variant (SNV) analysis.

Briefly, SNV metric files were created from the genotyped IDAT files as input for the CNV pipeline based on the GenoTools IDAT processing.^31^ For this analysis, the Genome Reference Consortium Human Build 38 was used as a reference genome.^32^ The input SNV metric files were split into interval-based windows to generate sample-level feature matrices for downstream analysis. We applied pretrained Long Short-term Memory models in CNV-Finder, which were trained separately for deletions and duplication, using features based on B Allele Frequency, Log R Ratio, and dosage statistics. For this, deletions were identified using a Log R Ratio of < -0.2, duplications with a Log R Ratio of > 0.2, and insertions with a B Allele Frequency of 0.15-0.35 or 0.65-0.85.^30^ The analysis focused on genes/regions of interest which commonly harbour PD CNVs, including *PARK7/DJ-1*, *PINK1*, *PRKN*, and *SNCA*. This included a default 250kb buffer region for *PARK7/DJ-1*, *PINK1*, and *PRKN* as well as a 1,000kb buffer region for *SNCA* to adequately capture the variations in this gene.^30^ The final output of CNV-Finder includes app-compatible files for visualization using an interactive web application, and results were visually inspected for structural variants.

### Validation of copy number variations

The identified CNVs were confirmed using MPLA. For this, DNA was extracted from a blood sample using established methods. The commercially available SALSA® MLPA® P051 Parkinson Mix-1 kit by MCR Holland (Amsterdam, The Netherlands; http://www.mlpa.com) was used for the MLPA analysis. The kit included probes for *PARK7/DJ-1*, *ATP13A2*, *PINK1*, *PRKN*, and *SNCA*. The analysis was conducted at the Unistel Medical Laboratories in Cape Town, South Africa, according to the manufacturer’s protocol. The MLPA assays were run with positive reference samples (with known CNVs) and wild-type controls, and the results were analysed using the Coffalyser.Net v.250317.1029 software.

## Results

### Bioinformatic analysis confirms the presence of putative pathogenic missense variants

As part of developing and optimizing the pathogenic variant workflow, we confirmed that the workflow was able to detect four previously reported pathogenic variants, which were present in 11 individuals in the South African PD Study Collection. Of the 13 individuals known to carry pathogenic variants, one failed QC due to high missingness, and another, despite passing QC, did not have their pathogenic variant genotyped. This reflects a common limitation of genotyping arrays, which often fail to capture rare pathogenic variants.

When looking at the subset of individuals without a known genetic cause for PD, 637 successfully passed QC. After filtering for PD phenotypes and using *in-silico* pathogenicity prediction tools, 69 variants were identified across nine chromosomes. After further clinical significance filtering to retain variants classified as “pathogenic” or “likely pathogenic”, 16 unique heterozygous variants in 47 individuals were prioritized. These were in the following seven genes: *GBA1* (n=18), *PRKN* (n=14), *LRRK2* (n=5), *PLA2G6* (n=5), *PINK1* (n=3), *POLG* (n=1), and *SYNJ1* (n=1). Sanger sequencing confirmed 15 of the 16 prioritized variants (**Table 1**). The variant rs191486604 (p.G430D) in *PRKN*, was not validated, suggesting a potential false-positive call in the initial workflow. The GBA1-PD Browser v1.0 classified the two *GBA1* variants as mild risk variants.^5^ Co-segregation analysis was performed for five families, where family member DNA was available. This included the following four variants: rs75548401 (p.T408M) and rs76763715 (p.N409S) in *GBA1* as well as rs199657839 (p.R334C) and rs149953814 (p.P437L) in *PRKN*. Several variants were detected in unaffected relatives, precluding co-segregation with PD. This observation is consistent with *PRKN*, where carriers may be heterozygous while probands are homozygous or compound heterozygous, and for *GBA1*, which is known to exhibit incomplete penetrance. The absence of symptoms among young carriers limits definitive conclusions, underscoring the need for continued clinical follow-up. Additionally, 18 VUS were identified in 27 unrelated cases in the following genes (**Supplementary Table 7**): *SYNJ1* (n=8), *PINK1* (n=6), *PRKN* (n=4), *GBA1* (n=3), *FBXO7* (n=2), *LRRK2* (n=2), *HTRA2* (n=1), and *PARK7/DJ-1* (n=1). The GBA1-PD Browser v1.0 classified the *GBA1* VUS variants as unknown severity.^5^

**Table 1:** Single nucleotide variants identified with the computational workflow which are classified as pathogenic, likely pathogenic, or conflicting pathogenicity.

| GP2 sample ID/<br>our lab ID | Gene | Variant (Amino acid<br>change/ rsID) | Family history of<br>PD | EOPD/LOPD |
| --- | --- | --- | --- | --- |
| 000163/ 10.111 | GBA1 | p.T408M / rs75548401 | No | EOPD |
| 000286/ 95.41 | GBA1 | p.T408M / rs75548401 | No | LOPD |
| 000339/ 96.91 | GBA1 | p.T408M / rs75548401 | Yes | LOPD |
| 000355/ 97.37 | GBA1 | p.T408M / rs75548401 | No | LOPD |
| 000510/ 12.797 | GBA1 | p.T408M / rs75548401 | Yes | LOPD |
| 000633/ 13.509 | GBA1 | p.T408M / rs75548401 | Yes | LOPD |
| 000066/ 67.7 | *GBA1* | p.T408M / rs75548401 | No | LOPD |
| 000113/ 81.61 | *GBA1* | p.T408M / rs75548401 | No | LOPD |
| 000133/ 82.45 | *GBA1* | p.N409S / rs76763715 | No | LOPD |
| 000176/ 83.41 | *GBA1* | p.N409S / rs76763715 | No | EOPD |
| 000206/ 88.29 | *GBA1* | p.N409S / rs76763715 | No | LOPD |
| 000357/ 97.43 | *GBA1* | p.N409S / rs76763715 | Yes | LOPD |
| 000417/ 10.327 | *GBA1* | p.N409S / rs76763715 | No | LOPD |
| 000427/ 11.59 | *GBA1* | p.N409S / rs76763715 | No | LOPD |
| 000580/ 13.266 | *GBA1* | p.N409S / rs76763715 | No | EOPD |
| 000599/ 13.352 | *GBA1* | p.N409S / rs76763715 | Yes | LOPD |
| 000273/ 12.87 | *GBA1* | p.N409S / rs76763715 | No | EOPD |
| 000146/ 82.91 | *GBA1* | p.N409S / rs76763715 | No | LOPD |
| 000437/ 12.705 | *LRKK2* | p.L119P / rs33995463 | No | LOPD |
| 000195/ 99.48 | *LRKK2* | p.R793M / rs35173587 | No | LOPD |
| 000366/ 98.33 | *LRKK2* | p.R793M / rs35173587 | No | LOPD |
| 000154/ 83.00 | *LRKK2* | p.R793M / rs35173587 | No | LOPD |
| 000545/ 12.925 | *LRKK2* | p.R1325Q / rs72546338 | No | LOPD |
| 000293/ 95.66 | *PINK1* | p.A339T / rs55831733 | No | LOPD |
| 000020/ 55.45 | *PINK1* | p.A359T / rs76753586 | No | LOPD |
| 000052/ 65.68 | *PINK1* | p.A494V / rs542258150 | No | LOPD |
| 000107/ 81.06 | *PINK1* | p.A494V / rs542258150 | No | LOPD |
| 000248/ 94.58 | *PLA2G6* | p.S34L / rs147948449 | Yes | LOPD |
| 000458/ 12.061 | *PLA2G6* | p.S34L / rs147948449 | No | LOPD |
| 000623/ 13.47 | *PLA2G6* | p.D31N / rs150024227 | No | LOPD |
| 000050/ 65.26 | *PLA2G6* | p.G361S / rs143826762 | No | LOPD |
| 000074/ 11.849 | *PLA2G6* | p.G361S / rs143826762 | No | LOPD |
| 000236/ 92.97 | *POLG* | p.V1044A / rs150233690 | No | LOPD |
| 000289/ 95.44 | *PRKN* | p.R334C / rs199657839 | No | LOPD |
| 000194/ 84.3 | *PRKN* | p.R334C / rs199657839 | No | LOPD |
| 000628/ 13.497 | *PRKN* | p.R334C / rs199657839 | No | EOPD |
| 000405/ 10.306 | *PRKN* | p.R334C / rs199657839 | No | EOPD |
| 000552/ 12.964 | *PRKN* | p.R334C / rs199657839 | No | EOPD |
| 000578/ 13.228 | *PRKN* | p.R334C / rs199657839 | No | LOPD |
| 000582/ 13.268 | *PRKN* | p.R334C / rs199657839 | No | LOPD |
| 000616/ 13.429 | *PRKN* | p.R402C / rs55830907 | No | EOPD |
| 000375/ 99.49 | *PRKN* | p.R402C / rs55830907 | Yes | LOPD |
| 000478/ 12.242 | *PRKN* | p.R402C / rs55830907 | No | EOPD |
| 000195/ 99.48 | *PRKN* | p.R402C / rs55830907 | No | LOPD |
| 000447/ 11.833 | *PRKN* | p.P437L / rs149953814 | No | LOPD |
| 000050/ 65.26 | *PRKN* | p.P437L / rs149953814 | No | LOPD |
| 000060/ 12.858 | *SYNJ1* | p.I707T / rs147929290 | Yes | LOPD |

This table provides the variants of interest classified as “pathogenic”, “likely pathogenic”, or “conflicting pathogenicity” as of July 2025 defined by ClinVar. EOPD, early-onset PD; LOPD, late-onset PD; PD, Parkinson’s disease.

### Copy number variant analysis reveals pathogenic structural arrangements

The CNV analysis using CNV-Finder identified 18 exonic CNVs in *PRKN* (n=15) and *SNCA* (n=3) in PD probands, along with an additional *SNCA* CNV of interest in the 5’ untranslated region in one proband (**Table 2** and **Supplementary Table 6**). The majority of the CNVs were located in *PRKN*, where ten *PRKN* CNVs were previously identified in probands and two in family members in the study collection.^15,33–35^ The new *PRKN* CNV findings include one duplication and four deletions, all of which were confirmed using MLPA, where three are homozygous. Representative examples of the graphical results from CNV-Finder are provided in **Figure 2A and B**. Of the 12 exons in *PRKN*, the CNVs identified were localised to exons 2 to 6. **Supplementary Figure 1** provides example visualizations of a heterozygous and a homozygous CNV in exon 4 (**Supplementary Figure 1**). The five cases carrying newly identified *PRKN* CNVs had no PD family history and an age of onset which ranged from 12-45 years.

**Figure 2:**
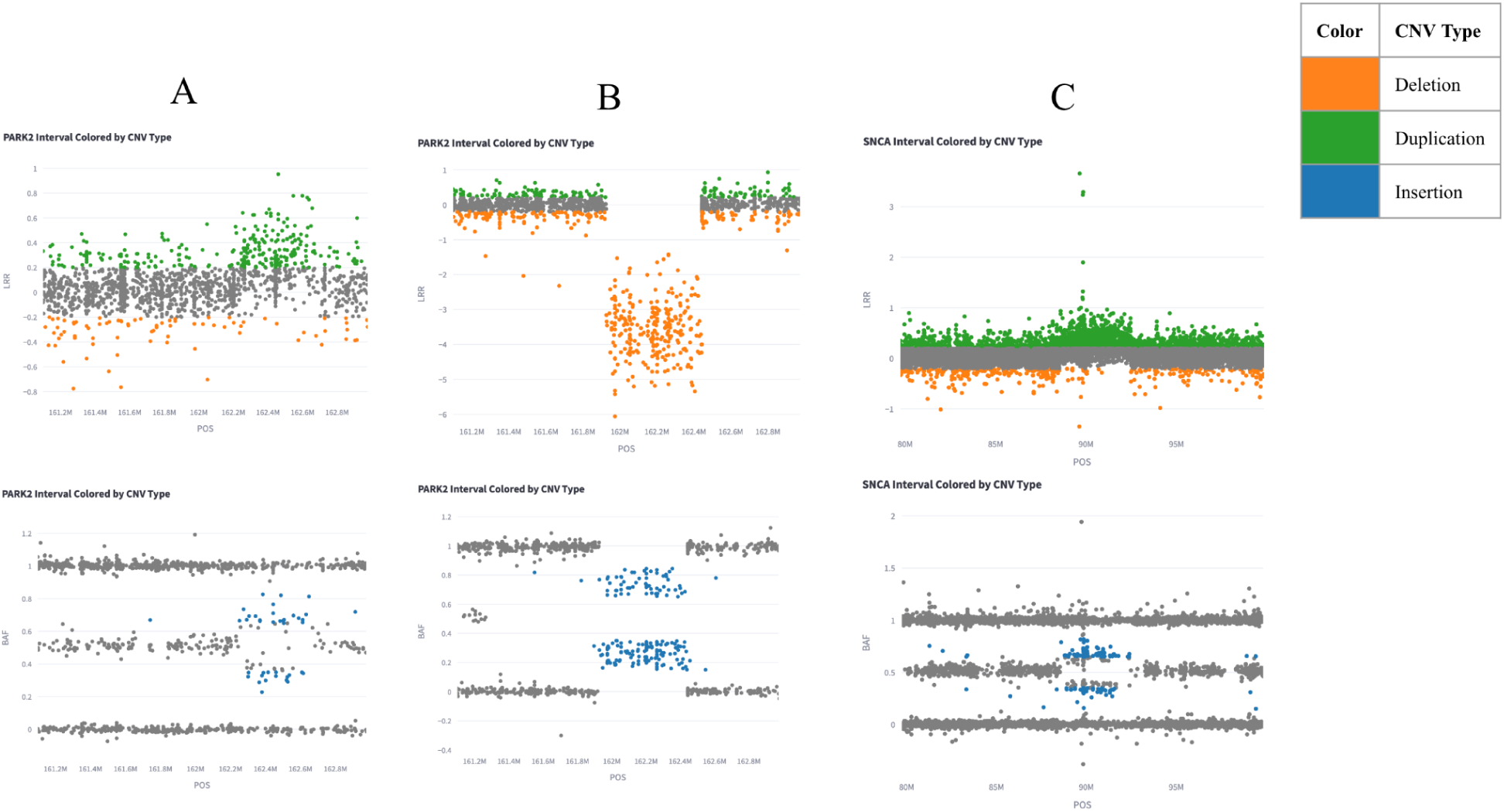
Representative CNV-Finder results showing (A) PRKN duplication, (B) PRKN deletion, and (C) SNCA whole gene duplication. CNVs are determined using visual inspection, prediction values, and the following ranges: Deletions: Log R Ratio (LRR) < −0.2, Duplications: LRR > 0.2, Insertions: 0.15 < B Allele Frequency (BAF) < 0.35 or 0.65 < BAF < 0.85.

**Table 2:**
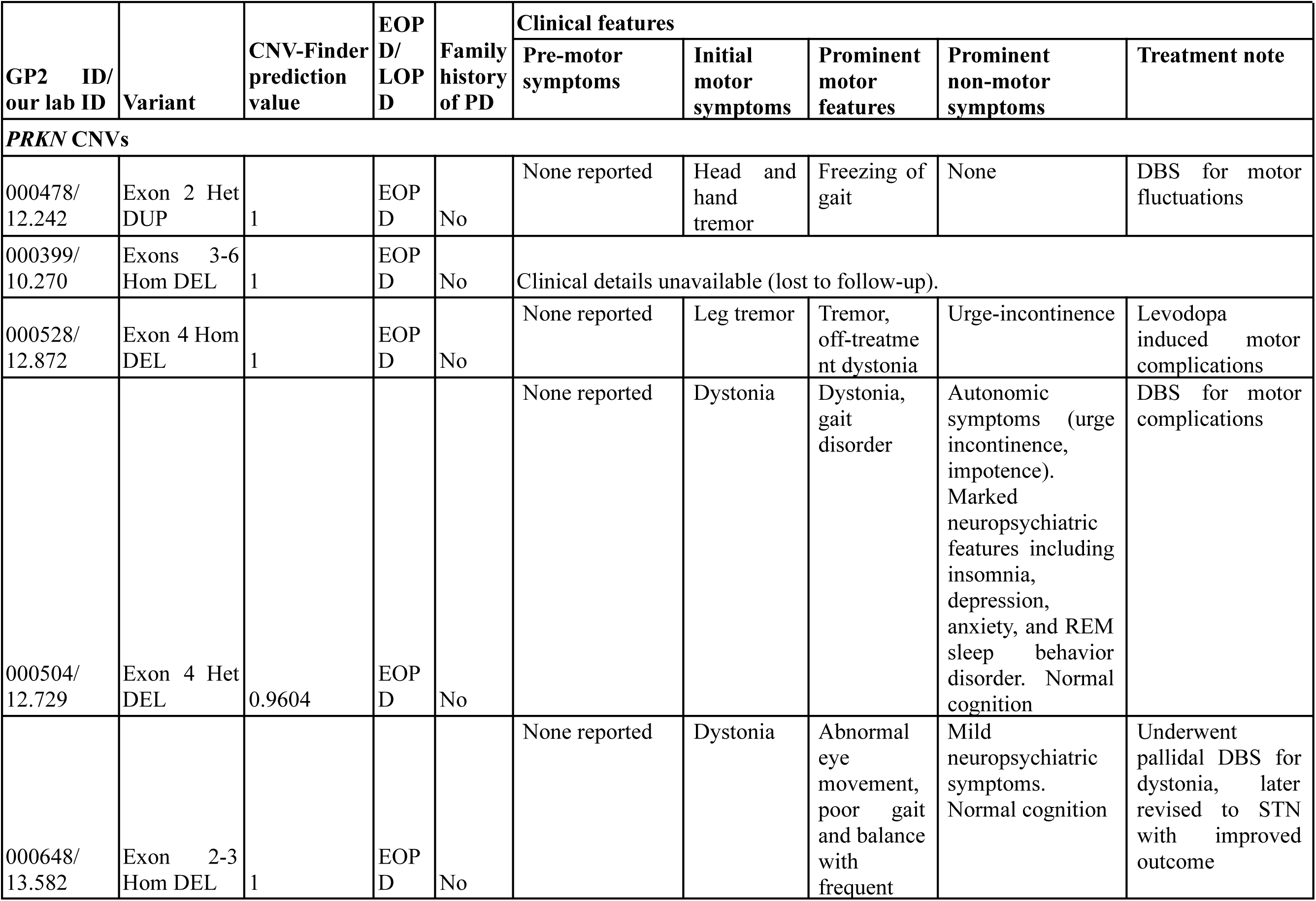
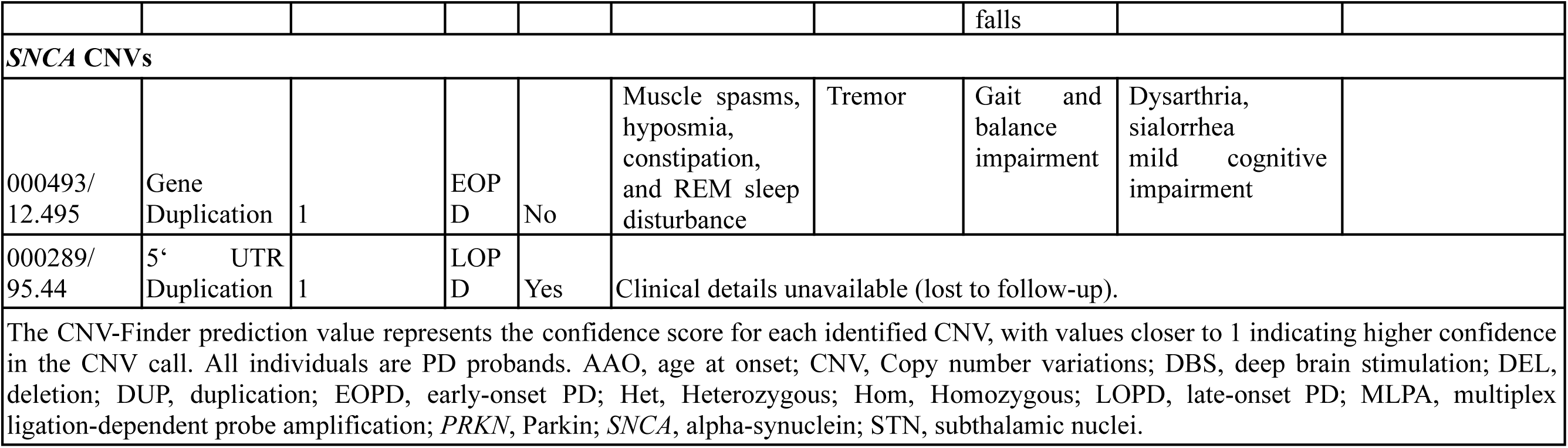
Novel prioritized copy number variations identified in the present study.

For *SNCA*, five whole gene multiplications that were previously identified in two families were all confirmed with CNV-Finder. This included three individuals with gene duplications in one family and two individuals with triplications in another.^33,36^ Moreover, one new *SNCA* whole gene duplication was identified using CNV-Finder (**Figure 2C**) and validated with MLPA. This individual has an age at onset in their 20s and no family history of PD. Additionally, a CNV was identified in the 5’ untranslated region of *SNCA* in one case with an age at onset in their 60s and a family history of PD (**Supplementary Figure 2**). However, due to the intronic location of this CNV, confirmation with MLPA was not possible.

Detailed clinical information for each novel case is presented in **Table 2**. We also observed *PRKN* cases with two CNVs that might indicate biallelic variation. These potential pairs were already known from earlier screening efforts, but the second CNV in each case fell below the CNV-Finder prediction score threshold and was excluded, as indicated in **Supplementary Table 6**. When considering both the pathogenic SNV and CNV analysis, there were no new compound heterozygous variants in the recessive genes detected in the South African PD Study Collection.

## Discussion

This study is the largest genetic screening for PD pathogenic missense and CNVs completed to date for the South African PD Study Collection. We successfully identified 16 missense variants, 18 VUS, and seven CNVs. Additionally, the new findings were confirmed using wet-laboratory approaches including Sanger sequencing and MLPA. The *PRKN* p.G430D variant did not validate with Sanger sequencing, suggesting it may represent a false positive. In contrast, the intronic *SNCA* CNV could not be assessed because the MLPA kit does not include probes covering this region, preventing experimental confirmation.

Integrating the findings from this study with previous screening efforts by the South African PD Research Group, 12.63% of probands (n=87) were found to carry a pathogenic variant across nine genes (**Table 3**). When considering only confirmed variants, excluding those with incomplete compound heterozygous findings or variants of uncertain penetrance, 3.48% of probands (n=24) harbored a variant that could fully explain their genetic basis for PD. These frequencies are notably lower than those reported in other global cohorts, where pathogenic variants have been detected in upwards of 20% of PD cases, highlighting potential population-specific differences in genetic risk.^1,6,37,38^

**Table 3:**
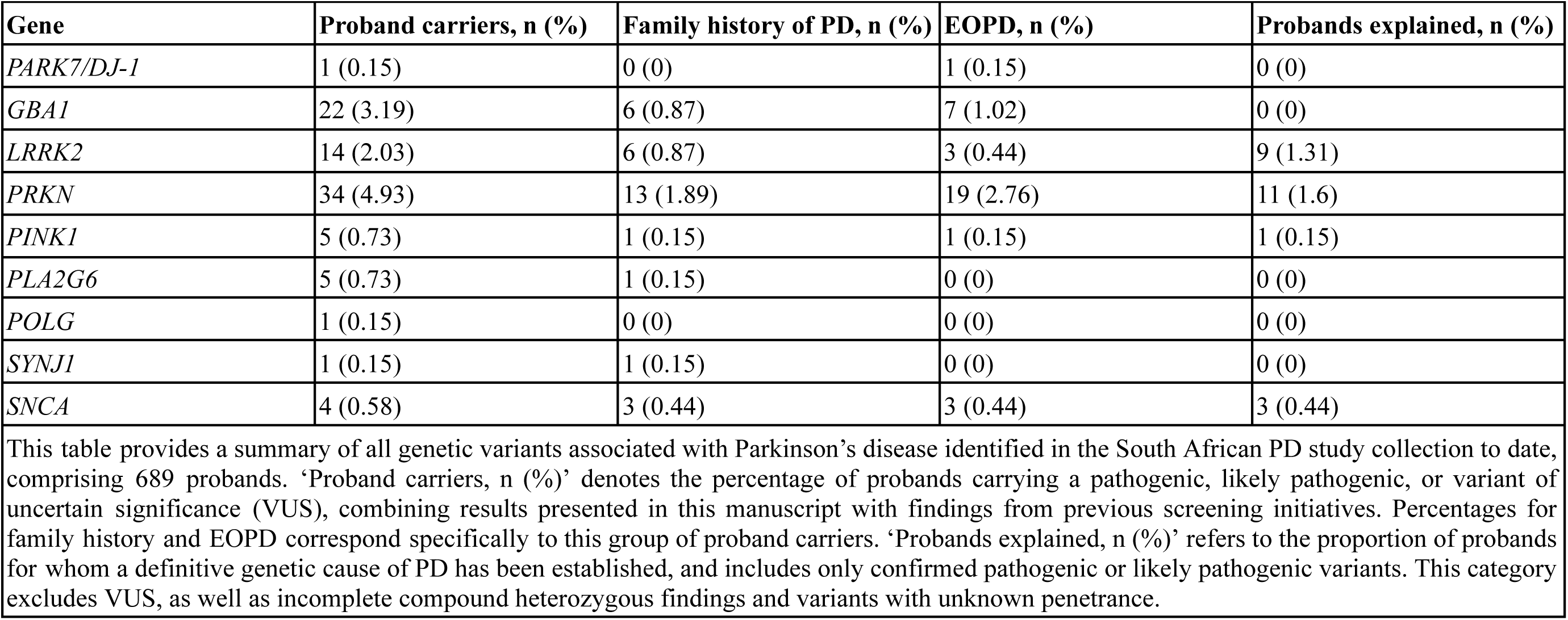
Overview of the variants identified in study collection probands.

In total, the presence of 16 putative pathogenic missense variants were confirmed in 47 individuals with PD. Several well-established PD-associated genes, including *GBA1*, *LRRK2*, *PINK1*, *PRKN*, *PLA2G6*, *POLG*, and *SYNJ1*, were found to harbor variants in our study collection. The most frequent were *GBA1* variants rs75548401 (p.T408M) and rs76763715 (p.N409S), which are known to have higher frequencies in European populations compared to African populations.^39^ Rare variants in *LRRK2* and *PINK1* were also identified, several of which have been proposed as familial causes of PD and are predicted to alter kinase function.^40,41^ Variants in *PRKN*, *PLA2G6*, *POLG*, and *SYNJ1* were observed at low frequencies, with *in-silico* tools supporting pathogenic classifications. This study made use of raw (non-imputed) genotyping data, which likely contributed to the limited number of pathogenic rare variants detected. Our analysis identified 18 VUS, assessed using ClinVar, the clinical implications of which remain to be established. Notably, the majority of these variants have been previously reported to be associated with PD in African, Asian, and European populations,^42–44^ suggesting potential relevance that warrants further investigation. Collectively, these findings highlight genetic contributors to PD in the South African cohort and emphasize the importance of variant characterization in underrepresented populations.

Furthermore, our findings highlight both previously reported and novel CNVs within our study collection. Notably, all five newly identified *PRKN* CNVs carriers presented with early-onset PD (<50 years) (**Table 2**), consistent with prior reports.^12,45^ All five cases reported no family history for PD. In terms of clinical manifestations, three of the *PRKN* CNV carriers displayed features consistent with *PRKN*-associated PD, albeit with variable presentations. Age at onset ranged from 12 to 45 years. Two presented with dystonia at onset, causing diagnostic delays, including one initially misdiagnosed as dopa-responsive dystonia. Another showed a tremor-dominant phenotype with typical progression, while autonomic symptoms such as urge incontinence and impotence were noted in two cases. Mild neuropsychiatric manifestations, including depression, anxiety, and REM sleep behaviour disorder, occurred in three individuals. Notably, three underwent deep brain stimulation, all with favourable outcomes. Overall, these findings reflect the phenotypic heterogeneity of *PRKN*-associated PD, although individuals still present with the characteristic features of early-onset, good levodopa response, and preserved cognition.^46^

The genomic distribution of *PRKN* CNVs in our dataset further supports observations in literature,^8,45,47^ with most occurring between exons 2 and 6, with exon 4 being the most frequently affected region. Importantly, *PRKN* is well known for harboring compound heterozygous variants, with most PD cases resulting from either two CNVs or a CNV combined with a pathogenic SNV. However, in the present study, no new compound heterozygous cases in *PRKN* were identified. Previous studies have suggested that single CNVs in *PRKN* may increase PD risk,^48,49^ although this association remains inconclusive given contradictory findings in literature.^50,51^ In our dataset, two *PRKN* CNVs were observed as single events, with no accompanying pathogenic SNVs or second CNVs. This suggests that, for those individuals, *PRKN* CNVs did not contribute to PD through the typical biallelic mechanism described in the literature. It remains possible that these individuals carry more complex or hidden structural variants in *PRKN* that are not detectable with genotyping array data. Recent long-read sequencing studies have demonstrated that such approaches can resolve previously unexplained single-heterozygous *PRKN* cases by uncovering intricate structural alternations.^52^ However, in three probands, we identified and validated homozygous *PRKN* CNVs, providing strong evidence that these biallelic events are the genetic cause of their PD.

For *SNCA*, seven CNVs were identified in probands and family members with two being novel findings (**Table 2**). The individual with the gene duplication showed an early age at onset and no family history of PD. While CNVs in *SNCA* tend to follow an autosomal dominant inheritance pattern,^9^ this type of CNV has been observed in both familial and sporadic PD cases.^53,54^ This individual experienced a range of non-motor symptoms including muscle spasms, loss of smell, sleep disturbances, and constipation. Additionally, motor symptoms included resting tremor which started on the right side and moved to the left, weakened hand strength, as well as poor balance and coordination. The presence of the *SNCA* CNV provides evidence that this is likely the genetic cause of PD for this proband. Finally, they also reported the onset and progression of short term memory loss. While whole gene multiplications are the most common CNV for *SNCA*, duplications are more frequently observed in sporadic PD cases with less severe disease symptoms in comparison to triplications.^55,56^ The *SNCA* CNV identified in the 5’ untranslated region is of particular interest, as disruptions in this region may affect gene expression by altering regulatory elements and transcription factor binding.^51,57^ This individual has late-onset PD and a positive family history, with an affected maternal grandmother. No further clinical information is available as this individual was lost to follow-up. Given the limitations of array-based CNV calling, the presence of this variant would ideally require confirmation with independent validation methods; however, this was not possible as the CNV lies outside the *SNCA* coding region, which is not covered by MLPA.

The reported presence of PD-associated CNVs in individuals of African ancestry, particularly from sub-Saharan Africa, remains limited, although it is unclear whether this reflects small sample size, true rarity, or both. Consistent with this uncertainty, previous screening efforts identified only five probands with *PRKN* CNVs and no *SNCA* CNVs reported in individuals of African ancestry.^8^ Similarly, a recent study successfully identified *PRKN* CNVs in individuals with African and African admixed ancestry on a global scale, but with no *SNCA* CNVs detected.^58^ However, the reported frequency of *PRKN* CNVs in that study was 0.7% of cases, highlighting the low frequency of CNVs in African populations compared to other ancestral groups, where frequencies as high as 10.8% have been reported.^59^ Taken together, this available literature suggests low observed CNV frequencies in African populations and suggests that novel genetic contributors remain to be uncovered.

In our cohort, several individuals carried heterozygous *PINK1* or *PRKN* variants (either SNVs or CNVs) that, on their own, do not fulfil the expected autosomal recessive inheritance pattern typically associated with pathogenicity in these genes.^46,60^ Despite comprehensive screening using both SNV and CNV analyses, we did not identify second hits, for example additional coding variants or copy-number events. In the absence of confirmed compound heterozygosity or biallelic disruption, these cases remain unresolved. It is therefore plausible that some individuals may harbor additional pathogenic alterations that were not detectable with our current array-based CNV approaches.^52^

While a number of pathogenic variants and CNVs were prioritized in the analysis, the study has several limitations. Genotyping arrays are widely used for their cost-effectiveness and coverage of common variants, however, they have limited sensitivity for rare variant detection and do not capture novel pathogenic variants.^61^ Additionally, probe sensitivity varies across genotyping arrays, which can lead to the detection of false positives and false negatives, highlighting the importance of further validation. Moreover, the detection of true CNVs using genotyping arrays remains challenging. While various bioinformatics approaches are available for this analysis, there is considerable variability in these methods.^62^ Some compound heterozygous CNVs, particularly in *PRKN*, were missed because one of the CNVs in the pair received a low CNV-Finder prediction value and was therefore excluded. Furthermore, potential compound heterozygous CNVs occurring in trans across consecutive exons may also be missed without long-read sequencing or parental testing to resolve allele phase. This indicates a limitation of this tool currently and requires further refinement. The results from CNV detection must therefore be carefully evaluated, either by integrating multiple tools or through wet-lab confirmation. While this is possible for genes that are on MLPA kits, not all CNVs can be validated with this approach. Ultimately, long-read whole-genome sequencing or array comparative genomic hybridisation is required to confirm novel CNVs and complex structural arrangements, such as inversions, in this population. This includes variants in non-coding regulatory regions, such as the 5’ untranslated region of *SNCA*, which cannot be validated using MLPA, where preliminary evidence suggests this CNV may represent a true finding.

In conclusion, this study is the first large-scale, systematic SNV and CNV screening in Southern Africa. We identified and prioritized several pathogenic missense variants and CNVs, providing valuable insights into the genetic architecture of the disease in the South African population. While whole genome sequencing is increasing in popularity, the scalability and affordability of genotyping arrays secure their continued role in research contexts. Ultimately, ensuring that researchers maximize the utility of genotyping data is vital for large-scale pathogenic screening, especially in underrepresented populations where resources are limited. Notably, CNV-Finder performed well in our dataset, with MLPA confirming all prioritised CNVs, highlighting its utility as a rapid and cost-effective alternative to MLPA for large-scale CNV detection. Importantly, through our collaboration with the Global Parkinson’s Genetics Program (GP2), WGS data for our study collection will become available in the future, enabling a more comprehensive investigation of pathogenic SNVs and CNVs that may have been missed due to the inherent limitations of array-based analyses. Importantly, these findings indicate the limited number of pathogenic variants detected in the known PD genes. Consequently, this study provides an important foundation for future studies and highlights the need to include African populations in neurological disease research to advance our understanding of disease etiology.

## Data and code availability

The data was obtained from the Global Parkinson’s Genetics Program (GP2) and access can be requested through the Accelerating Medicines Partnership in Parkinson’s Disease (AMP-PD) via the online application process (https://www.amp-pd.org/). This analysis made use of CNV-Finder https://github.com/GP2code/CNV-Finder; DOI 10.5281/zenodo.14182563. Additionally, an overview of the analysis and any additional scripts can be found in the GP2 public domain on GitHub (https://github.com/GP2code/PD_genescreen_SouthAfr; DOI 10.5281/zenodo.17478293).

## Author contributions

K.S., L.M., and S.B. conceptualized the manuscript. K.S. and L.M. wrote the first manuscript draft. K.S., L.M., and N.K. analysed the data. L.M. performed some of the wet lab validation. J.C. and R.v.C. recruited the study participants and provided clinical information. S.B., I.F.M., and E.M. supervised the project. All authors reviewed, edited, and approved the final version of the manuscript for submission.

## Data Availability

https://github.com/GP2code/CNV-Finder

https://github.com/GP2code/PD_genescreen_SouthAfr

## Acknowledgements

We would like to acknowledge and thank the study participants for their contribution. This project was supported by the Global Parkinson’s Genetics Program (GP2; https://gp2.org). GP2 is funded by the Aligning Science Across Parkinson’s (ASAP) initiative and implemented by The Michael J. Fox Foundation for Parkinson’s Research (MJFF). For a complete list of GP2 members see https://doi.org/10.5281/zenodo.7904831. For the purpose of Open Access, the author has applied a CC BY public copyright licence to any Author Accepted Manuscript version arising from this submission. All figures were created using BioRender (https://www.biorender.com/). We thank Kate Andersh, Laurel Screven, and Kim Paquette for their work as scientific project managers for this project. We would like to acknowledge Jai Ramchandra for his assistance with the *GBA1* variant validation. We also acknowledge the Centre for High Performance Computing (CHPC, http://www.chpc.ac.za), South Africa, for providing computational resources. We acknowledge the Biostatistics Unit, Centre for Evidence-based Health Care, Tygerberg Campus, Stellenbosch University. *For open access, the author has applied a CC BY public copyright license to all Author Accepted Manuscripts arising from this submission*.

